# Patient Portal Enrollment, Use, and Care Utilization among Hospitalized Children

**DOI:** 10.64898/2026.01.09.26343715

**Authors:** Michael J. Luke, Yinlu Zhu, Courtney B. Wolk, Stephanie G. Menko, Danielle Capriola, Katherine Fuller, Philip V. Scribano, Christopher P. Bonafide, Aditi Vasan

## Abstract

Patient portals are designed to enhance patient and caregiver engagement by providing access to health information, communication with care teams, and telehealth services, yet their impact on health care utilization among hospitalized children is not well understood. In this cross-sectional study, we examined associations between patient portal enrollment and use and hospitalization outcomes among 40,377 children admitted to an urban quaternary or suburban tertiary pediatric hospital between 2022 and 2024. Portal activity was defined by enrollment and any use (login, messaging, or telehealth) within specified periods before or during hospitalization. Using multivariable regression models adjusted for sociodemographic factors, neighborhood opportunity, insurance type, language, medical complexity, hospital site, and year, we evaluated associations with critical care admission, hospital length of stay (LOS), and 30-day readmissions. Portal enrollment and use in the year prior to hospitalization were associated with significantly lower odds of critical care admission and shorter LOS, with average reductions of 1.56 days for enrollment and 1.21 days for use. Neither portal enrollment nor use was significantly associated with 30-day readmissions. Future research should explore any potential mechanisms facilitating lower acuity at admission and shorter hospitalizations for those engaged with digital health.

## Introduction

Patient portals enable patients and caregivers to view health information, communicate with their care team, and participate in telehealth visits, among other features.^1^ Portals aim to help families make informed health decisions and enhance access to outpatient care to improve health and reduce acute care needs. Portal use is associated with improved patient satisfaction, medication adherence, and quality of care.^2^ In adults, patient portal adoption positively impacts health outcomes, but the relationship between portal use and health care utilization remains unclear.^3^ Moreover, because portal enrollment does not reliably translate into use,^4^ analyses that incorporate portal use data may better capture impacts on health care utilization. To our knowledge, no prior studies have explored the association of patient portal enrollment and use with health care utilization outcomes among hospitalized children.

## Methods

In this cross-sectional study of hospitalized children, we examined associations between patient portal activity (enrollment and use) and (1) admission to a critical care service, (2) hospital length of stay (LOS), and (3) 30-day readmission following discharge. Our sample included unique patients admitted to either an urban quaternary or suburban tertiary children’s hospital between 2022-2024 and seen within this health system for primary or subspecialty care. For critical care admission and LOS, we defined portal activity as enrollment and use in the year prior to admission. For readmissions, we defined portal activity as enrollment by the time of discharge and use in year prior to the index admission or during hospitalization. Portal use was defined as any login, message, or telehealth encounter in the specified period. This study was deemed exempt by our Institutional Review Board.

### Analysis

We used multivariable regression to examine associations between portal enrollment/use and critical care admission, LOS, and readmissions, controlling for age, race/ethnicity (as proxies for shared experiences of discrimination), neighborhood opportunity (using Child Opportunity Index 3.0), public or commercial insurance, primary language, and medical complexity (using Complex Chronic Conditions 3.0), with hospital site and year fixed-effects. We used a logistic regression model for the binary outcomes of critical care admission and readmissions, and negative binomial regression models for the continuous outcome of LOS. Admitting service was included as an additional covariate in models examining LOS and readmissions. We used Stata 18.0 for analysis.

## Results

This study included 40,377 patients (Table 1). Patients with portal enrollment (OR 0.68, 95%CI 0.61-0.76) or use (OR 0.64, 95%CI 0.58-0.70) prior to hospitalization had decreased odds of admission to a critical care service (Figure 1). Patients with portal enrollment (IRR 0.68, 95%CI 0.63-0.74) and use (IRR 0.74, 95%CI 0.69-0.80) prior to hospitalization also had significantly shorter LOS, with an average reduction of 1.56 and 1.21 days, respectively. Neither portal enrollment (OR 1.17, 95%CI 0.93-1.49) nor use (OR 1.18, 95%CI 0.96-1.45) were associated with a significant change in odds of 30-day readmissions.

**Table 1:** Sample Characteristics.

| **Sample Characteristics** |  | **Overall** |
| --- | --- | --- |
|  |  | **N=40,377** |
| **Age Category** | < 13 (Caregiver Access) | 30,151 (75%) |
|  | >= 13 (Patient Access) | 10,226 (25%) |
| **Race and Ethnicity** | Asian | 1,772 (4%) |
|  | Non-Hispanic White | 18,816 (47%) |
|  | Hispanic or Latino | 5,224 (13%) |
|  | Non-Hispanic Black | 10,069 (25%) |
|  | American Indian or Alaskan Native | 24 (0%) |
|  | Native Hawaiian or Pacific Islander | 13 (0%) |
|  | Multi-Racial | 1,717 (4%) |
|  | Other | 2,257 (6%) |
|  | Unknown | 485 (1%) |
| **Insurance Coverage** | Public | 17,761 (44%) |
|  | Private | 22,567 (56%) |
|  | Other | 49 (0%) |
| **Complex Chronic Conditions** | 1+ | 18,553 (46%) |
| **Preferred Language** | English | 38,170 (95%) |
|  | Spanish | 1,304 (3%) |
|  | Other | 903 (2%) |
| **Child Opportunity Index** | Very Low | 7,640 (19%) |
|  | Low | 4,616 (11%) |
|  | Moderate | 4,709 (12%) |
|  | High | 7,793 (19%) |
|  | Very High | 15,619 (39%) |
| **Hospital Site** | Suburban Community Site | 7,783 (19%) |
|  | Urban Academic Site | 32,594 (81%) |
| **Pre-Admission Portal Use** | Enrollment | 37,404 (93%) |
|  | Use | 35,572 (88%) |
| **Average Unadjusted Rates of Outcomes of Interest** | Admissions to Critical Care | 3,942 (13%) |
|  | Length of Stay (days) | 4.1 |
|  | Readmissions | 2,721 (7%) |

**Figure 1.**
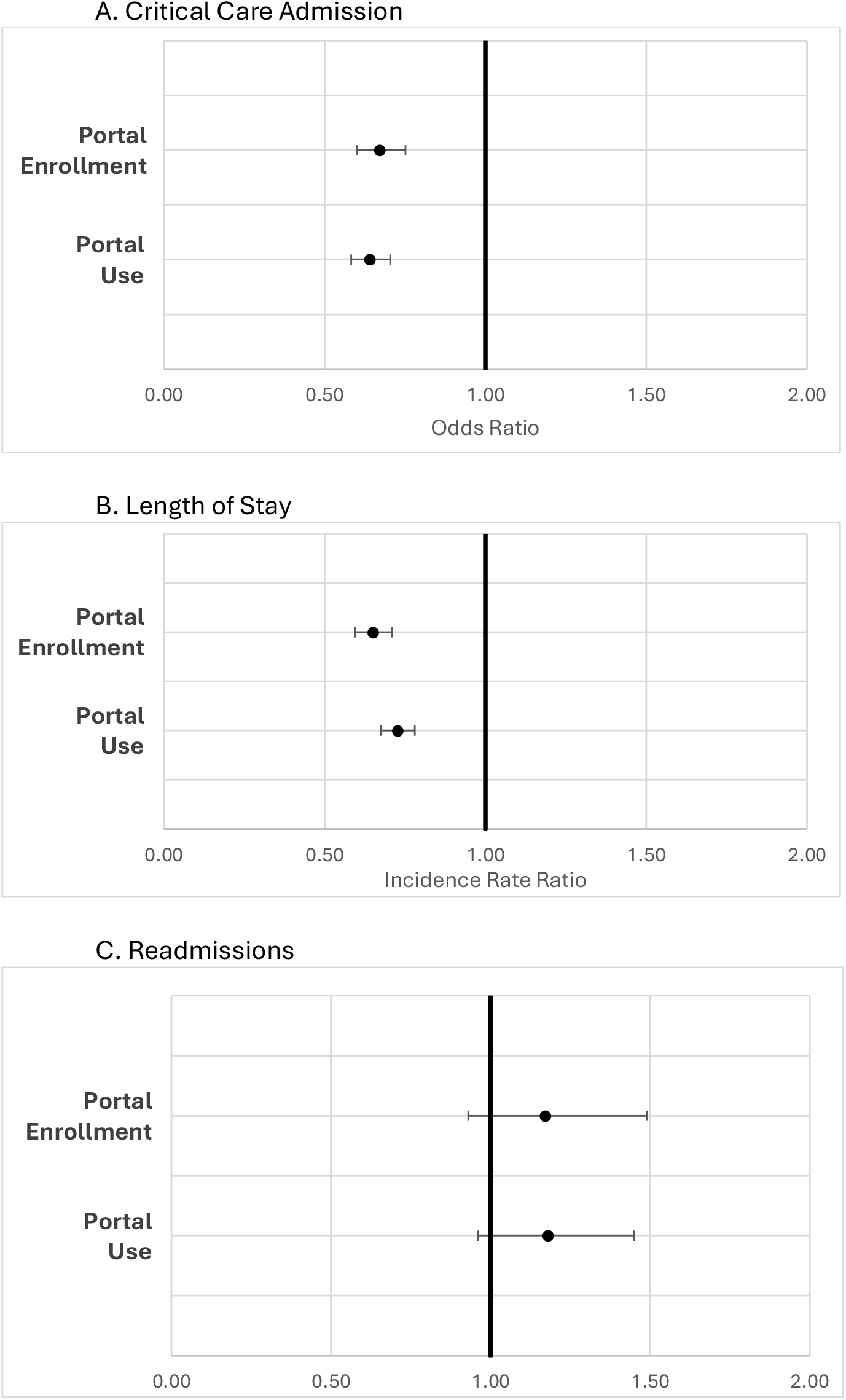
Association of Portal Enrollment and Use with Care Utilization.

## Discussion

Our findings suggest that patient portal enrollment and use are associated with decreased need for critical care admission and shorter average LOS. This association is likely multifactorial and could be partly mediated by healthier status at presentation, resulting from increased health care engagement. Prior data shows associations between portal enrollment and increased well visit attendance among infants,^5^ as well as reduced emergency department visits and hospitalizations among children with asthma.^6, 7^

We found no association between portal enrollment or use and 30-day readmissions. This is consistent with mixed findings in adult literature,^3^ and likely reflects the many complex factors that drive readmission rates. Portals may improve engagement with preventative care, including follow-up appointment attendance, but improved attendance at follow-up appointments could also lead to early detection of indications for readmission. Some post-discharge supports, like nurse telehealth follow-ups, have been linked to increased readmissions.^8^ Similarly, increased portal use may reflect increased concern from caregivers about their children’s health, leading them to seek acute care.^9^

While this study includes a large, diverse sample size across two hospitals, it has limitations. Our population exhibited higher levels of portal enrollment and use compared to other studies,^10^ limiting generalizability. We also lack granular data on the frequency of portal use and use of distinct portal features. Finally, there may be other confounding factors not accounted for in our analysis, and further research is needed to identify potential mechanisms for our findings.

## Data Availability

Interested parties can contact the first author for any data sharing requests.

